# AI-driven Mental Health Decision Support Enhances Clinician Resilience and Preparedness

**DOI:** 10.1101/2025.10.29.25338983

**Authors:** Margareta-Theodora Mircea, Jessica McFadyen, Ross Harper, Max Rollwage, Tobias U. Hauser

## Abstract

**Objectives:** Mental health services are facing unprecedented demand, placing significant pressure on clinicians to conduct timely and effective patient assessments. Rising staff turnover and burnout threatens service quality across many countries. This study examined whether providing clinical information, collected via an AI-enabled decision support tool for mental health assessments in the UK’s National Health Service (NHS), could improve clinician wellbeing and patient assessment performance.

**Method:** We surveyed mental health clinicians (N=131) from nine NHS Mental Health Talking Therapies services on how the information provided by an AI-based decision-support tool impacted their experience with conducting clinical assessments. Clinicians reported on assessments where information from the AI tool was available, as well as when it was not (e.g., GP referrals or telephone intakes). Outcomes included clinician wellbeing, task performance, and cognitive load during assessments, with additional analyses assessing the influence of moderating factors, such as clinician experience, workload, and exposure to the tool.

**Results:** Relative to traditional methods, assessments supported by information provided by the AI tool were associated with significantly higher clinician wellbeing and task performance, and significantly lower cognitive load, irrespective of the clinician’s experience. These benefits were magnified by workload.

**Conclusion:** These findings provide evidence that AI-powered pre-assessment tools can enhance clinician experience by improving wellbeing, boosting task performance, and reducing cognitive burden. By addressing systemic drivers of burnout, such tools offer a scalable intervention to support workforce sustainability and service quality in mental health care.

## Introduction

Mental health services are experiencing unprecedented demand, with dramatic increases of up to 40% compared to pre-pandemic levels (NHS England, 2024). This is further exacerbated through above-average staff turnover in mental health services and disproportionately high vacancy rates (Long et al., 2023). Due to budgetary constraints across many healthcare services worldwide, this directly translates to mounting pressures on mental health services, creating unsustainable workloads for clinicians.

Large caseloads and insufficient organisational support are well-established predictors of reduced wellbeing and heightened emotional exhaustion, the core features of burnout (Kim et al., 2018; Barnes, 2024). Burnout, in turn, directly compromises patient care: only 28.3% of patients treated by burnt-out therapists demonstrate meaningful improvement, compared with 36.8% of those treated by therapists without burnout (Sayer et al., 2024; Delgadillo et al., 2018; Salyers et al., 2017). Taken together, these findings highlight that safeguarding clinician wellbeing is not only essential for workforce sustainability but also for ensuring the effectiveness of psychological treatment.

A case example is the UK’s National Health Service (NHS). Within NHS Talking Therapies, Psychological Wellbeing Practitioners (PWPs) are particularly susceptible to burnout, with evidence indicating that elevated work demands substantially contribute to this risk (Westwood et al., 2017; Steel et al., 2015). PWP trainees appear especially vulnerable, reporting higher levels of stress and burnout than both their more experienced colleagues and the wider healthcare workforce (Owen et al., 2021). One contributing factor may be the relatively brief training period before trainees begin clinical practice, which can limit their knowledge and confidence in therapeutic delivery (West Suffolk NHS Foundation Trust, n.d.). As knowledge and confidence are recognised protective factors, this suggests that enhancing preparedness - during training or through other means - may help reduce burnout risk (Kim et al., 2018).

Here, we examined whether scalable tools that leverage artificial intelligence might boost clinician preparedness and mitigate risk factors of burnout. Virtual triage tools offer a scalable and effective solution to reducing the cognitive and emotional burden on therapists by supporting clinical tasks such as diagnoses, risk assessment, and treatment planning (Gellert et al., 2023). However, most existing systems are rule-based and operate within fixed protocols (Willman, 2023; Turner et al., 2021), limiting their ability to capture the complexity of patient presentations. A key promise of artificial intelligence (AI) is to expand such capabilities substantially. AI-driven inference engines can adapt dynamically to patient input, medical history, and risk factors, mirroring the flexible reasoning of human clinicians and providing substantially more clinical value (Rollwage et al., 2024b).

Here, we investigate an AI decision support tool called Limbic Access, a certified UK CE-marked Class IIa medical device that facilitates chatbot-based patient intake, initial clinician-led assessments, and overall decision support for clinical and administrative staff. This AI decision support tool interprets symptoms, evaluates risk, and prioritises relevant clinical needs with diagnostic accuracy that surpasses human assessment (Rollwage et al., 2024b). It has been shown to successfully increase patient access to healthcare (Habicht et al., 2024), streamline clinical workflows (Rollwage et al., 2023), increase efficiency of healthcare provision (Rollwage et al., 2024a), and improve patient recovery rates (Rollwage et al., 2024a; Rollwage et al., 2023).

In this study, we assess the impact of this decision support tool more specifically on clinician wellbeing and performance in conducting initial patient assessments. As this tool equips clinicians with patient information in advance of appointments, we hypothesised that it would ease the cognitive and emotional burden on the clinician performing the assessment (Gellert et al., 2023; Gilburt & Mallorie, 2024) and improve self-perceived task performance.

## Materials and methods

### Study design

We employed a within-subjects, comparative survey design to evaluate the impact of the AI decision support tool on clinician wellbeing, task performance and cognitive load. We asked PWPs from nine different NHS Talking Therapies services across England to participate. NHS Talking Therapies services (formerly known as Improving Access to Psychological Therapies, or IAPT) are a UK government initiative delivering evidence-based psychological interventions within a stepped-care model. They primarily serve adults experiencing common mental health difficulties such as anxiety disorders and depression and offer a range of talking therapies treatments (e.g., guided self-help, cognitive-behavioural therapy, counselling, and interpersonal therapy).

In an online survey, clinicians were asked to report their experiences with patient assessments in which they had been provided with clinical information from the AI decision support tool (available only when patients had self-referred through the tool) versus when this information had not been provided (for patients referring to the service through other means, such as referrals from their GP or self-referring over telephone). This within-subjects comparative approach, where clinicians reported on their experiences with both types of information sources, allowed each PWP to serve as their own control, thereby enhancing the sensitivity of our measurements to the differential effects of the tool-derived information.

### Participants

A cohort of 146 PWPs were recruited for the study. They were invited to participate by members of the study research team during routine team meetings held within each NHS service. During these meetings, an overview of the study was provided, and interested PWPs were given a direct link to the online survey. As an incentive, participants were entered into a prize draw for a £10 Amazon voucher. All participants provided informed consent online before commencing the survey. This study was approved by the NHS London - Surrey Borders Research Ethics Committee (22/PR/1424).

The primary inclusion criterion was being currently employed as a PWP and actively conducting initial patient assessments within one of the participating services. Exclusion criteria were: (a) not having conducted any initial patient assessments in the 30 days preceding survey completion (n=2), or (b) within the preceding 30 days, having *only* conducted assessments informed by the AI decision support tool, or conversely, *only* assessments informed by traditional methods. This latter criterion was essential to ensure participants could provide valid within-subjects comparisons based on recent experience with both types of information sources. Thirteen participants were excluded due to only having done assessments for patients with AI decision support tool information (n=8) or only assessments without this information (n=5), leaving a final sample of 131 participants.

## Materials

### AI Decision Support Tool

The tool evaluated in this study was Limbic Access (https://limbic.ai/access), a commercially available, AI-powered digital product designed to function as an intelligent decision support tool for mental health services (**Fig. 1**). It functions as a chatbot positioned on a mental health provider’s web page, where new patients can self-refer easily and discreetly. Patients interact with Limbic Access through a conversational user interface (chatbot) and input information via free text, buttons, and checkboxes. The chatbot uses machine learning to make clinical inference about the most likely diagnoses of a patient and the most relevant associated symptoms, which is used to intelligently guide the referral process and collect valuable clinical patient information that is then summarised and conveyed to the clinical team.

**Figure 1.**
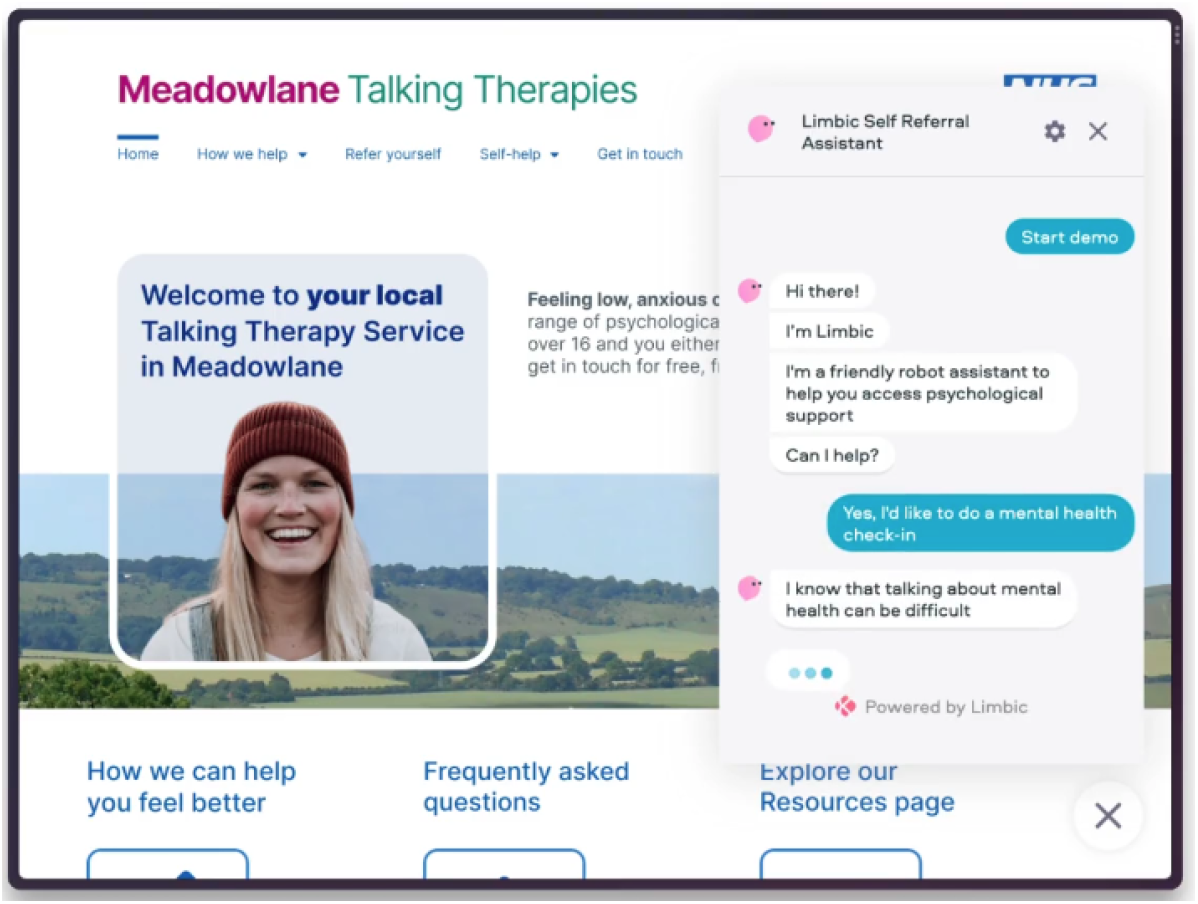
Conceptual mock-up of the AI-powered mental health chatbot as it might appear on a service’s landing page during patient self-referral.

For clinicians, the key feature is the structured clinical referral report that is automatically generated and integrated into the service’s electronic health record. Each report begins by clearly identifying it as a Limbic referral and includes the patient’s risk level. Patients that indicate high levels of acute risk (e.g., suicidal intent) are immediately signposted to emergency services and a “crisis alert” is sent to the service. The tool also performs eligibility checks, such as confirming the patient resides within the geographical catchment area of the specific NHS service. The core clinical information then follows in a structured sequence: the AI-inferred primary and secondary presenting problems, the patient’s stated expectations from support, “caseness” status (i.e., symptoms above a clinical threshold) across relevant diagnoses, and the condition the patient identified as their primary concern.

Standardised questionnaire - e.g., Patient Health Questionnaire-9 (PHQ-9; Kroenke et al., 2001) for depression and Generalized Anxiety Disorder-7 (GAD-7; Spitzer et al., 2006) for general anxiety - scores are presented to the clinician in an accessible format. A key intelligent feature of the AI decision support tool is the adaptive selection of additional symptom questionnaires for specific anxiety disorders (e.g., social anxiety, panic disorder), which are triggered by the AI during the patient referral process when clarification is needed to resolve uncertainty over the most likely presenting problems. This adaptive, personalised approach tailors the referral process to each patient by presenting only the most relevant questions, thereby improving efficiency and enhancing the accuracy of the inferred presenting problems provided to clinicians.

The report ends with the patient’s contact details and demographic information. This structured output provides clinicians with a comprehensive, pre-synthesised overview that extends beyond raw data capture. This clinical information which is automatically collected by the AI decision-support tool, enables the clinicians with a more comprehensive clinical profile of the patient before the clinical assessment which allows for a more in-depth preparation of the assessment. Rather than expending time and effort on information gathering, clinicians can direct their attention to higher-order tasks such as clinical reasoning, therapeutic engagement, and collaborative care planning.

### Traditional Referral Pathways

Comparator information sources included existing referral methods routinely used within the participating services, including self-referral (e.g., telephone call, static online forms on the service’s website, walk-ins), referrals from primary care (e.g., general practitioners), referrals from secondary care (e.g., specialist mental health services), and referrals from other agencies (e.g., social services). A critical distinction between these traditional referral pathways and the AI decision support tool is that the former typically provide less detailed and often unstructured patient information (e.g., basic demographic details, brief free-text descriptions, or checkbox answers), compared to the comprehensive, clinically-refined data gathered and processed by the AI decision support tool.

### Survey Instrument

A custom online survey was developed to capture participants’ comparative experience with patient assessments informed by the AI decision support tool versus those informed by information from traditional referral pathways. The survey first collected information about the participant’s professional role (e.g., job title, employment duration) and frequencies of patient assessments per week in the last 30 days. Participants then indicated the proportion of recent patient assessments for which they viewed information derived from the AI decision support tool (5-point Likert scale from “none” to “all”).

The core of the survey employed a structured comparative approach to assess key outcomes. These were administered in two distinct stages: first, an item set regarding assessments *with* the information provided by the AI decision support tool, covering clinician wellbeing, task performance and cognitive load (see **Outcome Measures**); second, the identical item set but instead based on experiences conducting assessments *without* the AI decision support tool-derived information (i.e., patient intakes through traditional referral methods).

## Outcome measures

The primary outcome measures were clinician wellbeing, task performance, and cognitive load, all assessed comparatively for experiences with assessments informed by the AI decision support tool versus those with information from traditional referral pathways. All questions used to assess the outcome measures are provided in the **Supplementary Material**.

### Clinician Wellbeing

Six items assessed clinicians’ affective states related to conducting assessments. These included both positive affect (feelings of energy, confidence, and comfort) and negative affect (stress, uncertainty, and anxiety). Each question was phrased as, “How [emotion] do you feel about these assessments?” where the emotion is the outcome measure (e.g., “stressed”) and “these assessments” refers to either those with AI decision support tool information provided or those without. Responses to all items were made on a 7-point Likert scale from “not at all” to “extremely”. Negative affect items were reverse scored, and all items were averaged to produce a single wellbeing measure.

### Task Performance

Seven items evaluated the perceived ease of performing key assessment-related tasks. These included preparing for the assessment, determining appropriate treatment, conducting a risk assessment, identifying a diagnosis, completing the assessment within the appointment time limit, building a patient relationship, and managing patient expectations. Each question was phrased as, “How easy is it to [task]?” where “task” is the variable (e.g., “complete an assessment”). Responses were made on a 7-point Likert scale from “not at all” to “extremely”, with responses averaged across items to produce a single task performance score.

### Cognitive Load

Cognitive load was measured using a modified version of the NASA Task Load Index (NASA-TLX; Hart & Staveland, 1988). The NASA-TLX has demonstrated strong internal consistency (e.g., Cronbach’s α = 0.80; Xiao et al., 2005) and moderate convergent validity with other workload measures (r = 0.33 to 0.49; Devos et al., 2020; Xiao et al., 2005).

Adaptations for this study involved removing the ‘physical demand’ subscale, as it was not relevant to the task, and omitting the pairwise weighting procedure for item pairs to reduce complexity. The modified scale comprised five items assessing: mental demand (“How mentally demanding is the assessment?”), hurried pace (“How hurried or rushed is the pace of the assessment?”), success in accomplishment (“How successful are you in accomplishing what you want to accomplish?”), effort (“How hard do you have to work to accomplish your level of performance?”), and negative affective response (e.g., insecurity, irritation, stress; “How insecure, discouraged, irritated, stressed, or annoyed do you feel?”). Responses were made on a 7-point Likert scale from “not at all” to “extremely” (adapted from the original 11-point scale to maintain consistency across all outcome measures administered in the survey), and the item “success in accomplishment” was reverse-scored. Responses were averaged across items to produce a single cognitive load score.

## Statistical analysis

Linear mixed-effects regression models were used to examine the impact of the AI decision support tool on clinician wellbeing, task performance and cognitive load, with each outcome predicted by a separate model. All analyses were conducted in Python (version 3.11) using the statsmodels package (version 0.14.3). Each clinician contributed data under two conditions (AI decision support tool-based assessments vs. traditional methods of referral-based assessments), resulting in 262 observations per model. A random intercept was included for each clinician. All continuous outcomes and predictors were standardised (z-scored) to allow effect size inferences from the resultant coefficients (β). To further explore item-level effects within each averaged outcome variable, paired t-tests were conducted for each specific questionnaire item, with p-values corrected for multiple comparisons using the false discovery rate (FDR) method.

In all models, we controlled for three potentially moderating variables: *experience* (employment duration), *workload* (number of patient assessments per week) and *exposure* (proportion of assessments in which clinicians received referral information from the AI decision support tool). Employment duration was included as an ordinal variable to determine whether the tool was equally effective for early-career and more experienced clinicians. The average number of patient assessments per week was used as a continuous variable, indicating workload intensity. Finally, the reported proportion of assessments in which clinicians received AI decision support tool referral information was included as an ordinal variable (“some”, “half”, or “most” assessments per week) to assess whether the frequency of exposure influenced perceived benefits.

For each of the three outcome variables, four models were estimated, yielding a total of twelve models. The first model included the main effects of the AI decision support tool and all three covariates (experience, workload, and exposure). The remaining three models each included an interaction term between the AI tool variable and one of the covariates, while controlling for the remaining covariates. This approach allowed us to test whether the impact of the AI decision support tool varied as a function of each covariate, while minimising multicollinearity among predictors.

## Results

We conducted a within-subjects comparative survey with 131 PWPs from nine NHS Talking Therapies services to assess how an AI decision support tool influenced clinician wellbeing, task performance, and cognitive load, while accounting for clinician experience, workload, and exposure to the tool.

### Clinician Characteristics

All 131 participants were PWPs (91.6% full-time) with a range of experience levels and seniority. Professional tenure was skewed toward early-career practitioners, with 90.8% being in post for under five years, including 1.5% for 1–3 months, 7.6% for 3–6 months, 15.3% for 6–12 months, 32.1% for 1–2 years, and 34.4% for 2–5 years. A smaller group had more than five years of experience (6.1% for 5–10 years, 3.0% for over 10 years). Clinicians reported conducting an average of 8.47 client assessments per week (SD = 5.3, range = 1 to 36).

Over half of clinicians (58.8%) reported that most of their assessments were supported by the AI decision support tool, and nearly a quarter (26.7%) reported roughly an equal split between referrals with and without AI tool information. Only 14.5% of clinicians reported that just some of their assessments were guided by the AI tool output. These varied usage figures provide a solid basis for evaluating the impact of the AI decision support tool in routine practice.

### AI Decision Support Tool Improves Clinician Wellbeing

Clinician wellbeing is closely tied to burnout risk and service quality (Sayer et al., 2024; Roy et al., 2020), and so we first examined whether the AI tool improved clinicians’ wellbeing during assessments. Clinicians were asked to report their energy levels, confidence, comfort, stress, uncertainty and anxiety associated with clinical assessments. Clinicians reported significantly higher wellbeing overall (**Fig. 2a**) when using the AI decision support tool (M = 4.7, SD = 0.9, on a scale from 0 to 6) compared with traditional referral information (M = 3.8, SD = 1.1; β = 0.77, 95% CI = [0.57, 0.97], P = 2.32 × 10^-14^). This reflects a large effect, with wellbeing scores approximately 15% higher when clinicians used the AI tool. This AI-enabled wellbeing boost was seen across all items: increased energy, confidence, and comfort, alongside decreased stressed, uncertainty, and anxiety (all pairwise t-tests P_FWE_ < .002) Out of the control variables, only “exposure” was a significant moderator, where clinicians who performed relatively more assessments using the AI decision support tool were even more likely to experience improved wellbeing for these assessments relative to those for patients coming through traditional referral methods (β = 0.34, 95% CI = [0.15, 0.52], P = 5.15 × 10^-4^). This suggests that the more often clinicians viewed AI-generated referral information in their routine work, the more beneficial this information was for their occupational wellbeing.

**Figure 2.**
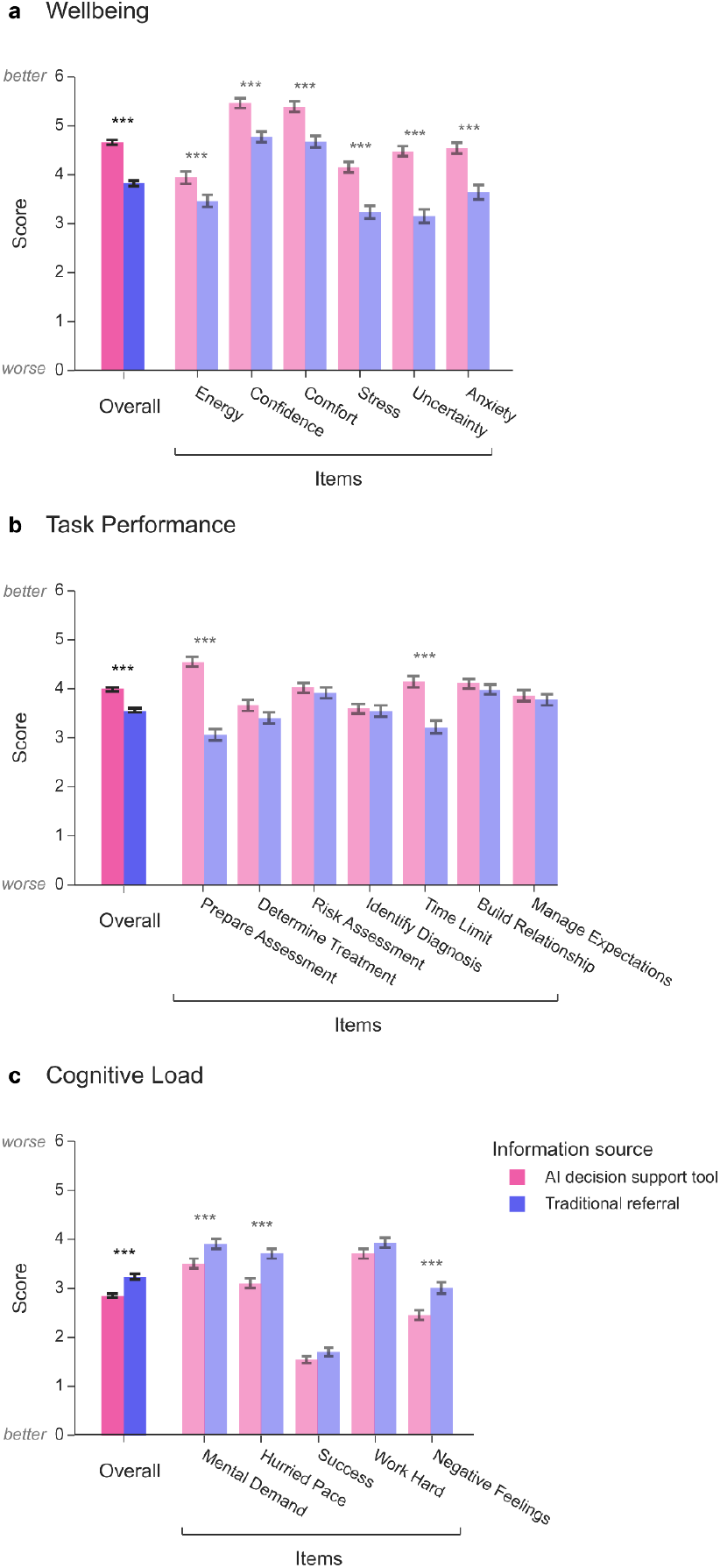
Clinician wellbeing, task performance, and cognitive load outcomes. **a**, Average wellbeing scores reported by clinicians for patient assessments that had information provided by the AI decision support tool (pink) or for those that had information from traditional referral methods (e.g., GP referrals, telephone intakes; purple). The overall score represents the average of each individual item. Error bars indicate standard error of the mean. Significance indicators for “overall” represents the regression coefficient from a linear mixed effects model for the information source predictor. Significance indicators for each individual item represent the family-wise error corrected results from a series of paired t-tests. * P < .05, ** P < .01, *** P < .001. **b**, Same as (**a**) except for task performance relating to the patient assessment. **c**, Same as (**a**) except for cognitive load (adapted NASA Task Index), with lower scores indicating reduced (and thereby improved) cognitive load.

### Enhanced Task Performance During Assessments

As effective assessment underpins accurate diagnosis and treatment planning, we next tested whether the AI tool enhanced clinicians’ performance during initial assessments. Clinicians rated the ease with which they prepared for assessments, determined treatment pathways, assessed risk, identified diagnoses, adhered to time limits, built relationships with the patient and managed their expectations. Results showed significantly higher task performance scores (**Fig. 5b**) when reporting on assessments supported by the AI tool (M = 4.6, SD = 1.0, on a scale from 0 to 6) compared to traditional referral information (M = 4.0, SD = 1.2), β = 0.42, 95% CI = [0.19, 0.66], p = 3.427 × 10^-4^). This indicates that the AI tool significantly improved perceived task performance, with a moderate effect size. Follow-up paired t-tests revealed this was driven predominantly by feeling more prepared for the assessment (P_FWE_ = 4.10 × 10^-13^) and being more able to complete the assessment within the appointment time (P_FWE_ = 7.67 × 10^-9^).

This enhanced task performance, made possible by the AI-generated reports, were accentuated for clinicians with a higher assessment workload (β = 0.26, 95% CI = [0.03, 0.49], P = 0.026) and for those exposed more to the AI reports (β = 0.36, 95% CI = [0.13, 0.59], P = 0.002). Employment duration of the clinicians did not significantly modulate the AI effects (β = -0.23, 95% CI = [-0.46, 0.01], P = 0.063), suggesting similar benefits of the AI decision support tool across clinician experience levels.

### Reduced Cognitive Load Through AI Assistance

Lastly, we assessed whether the AI tool reduced perceived cognitive demands, given the potential for cognitive load to compromise decision-making and increase stress. We measured cognitive load using a modified version of the NASA Task Load Index (NASA-TLX; Hart & Staveland, 1988) comprising five items assessing: mental demand, hurried pace, success in accomplishment, effort, and negative affective response.

Clinicians reported significantly reduced cognitive load when using the AI decision support tool (M = 3.5, SD = 0.9, on a scale from 0 to 6) compared with traditional referral information (M = 3.9, SD = 1.0), β = -0.48, 95% CI = [-0.65, -0.31], P = 2.955 × 10^-8^), with a moderate-to-large effect size (**Fig. 5c**). Follow-up paired t-tests on each item revealed that the AI tool was most effective at reducing mental demand, hurried pace, and negative feelings about the assessment (all P_FWE_ < 3.93 × 10^-4^), while there were no significant difference in the feelings of success or accomplishment, nor working hard, after family-wise error-correction (all P_FWE_ > .182). These findings suggest that the AI tool markedly reduced the cognitive demands of conducting patient assessments across multiple dimensions. These beneficial reductions in cognitive load were seen more for clinicians who conducted relatively more assessments where an AI-generate report was available (β = -0.18, 95% CI = [-0.35, -0.02], P = 0.032).

## Discussion

This study examined the impact of a clinical decision support tool for mental health assessments on clinicians’ everyday practice. We found that use of the AI decision support tool significantly improved clinician wellbeing and task performance, while also reducing the cognitive load associated with conducting clinical assessments. Importantly, these benefits were evident across varying levels of clinical experience and were particularly pronounced among clinicians with heavier assessment workloads. Overall, these findings suggest that deploying AI tools in clinical settings may not only enhance patient outcomes but also help clinicians optimise their daily performance. In the context of resource-constrained healthcare systems and increasing clinician strain, our results indicate that AI-enabled decision support tools could meaningfully improve clinician experience in domains closely linked to burnout and service quality.

Clinicians reported significantly better wellbeing when conducting patient assessments with information provided by the AI decision support tool compared to when such information was absent. Typically, clinicians begin assessments with very limited knowledge about a patient’s presenting problems, severity, or risk, which can heighten stress during initial consultations. In contrast, the AI-enabled decision support tool offers access to this information in advance, reducing uncertainty and potentially enhancing clinician wellbeing. Given that clinician wellbeing is a strong predictor of burnout risk (Meredith et al., 2022; Ashton-James et al., 2021; Roy et al., 2020), these findings suggest that AI-powered decision support tools may play a preventative role in mitigating burnout and its downstream effects on service quality and patient outcomes (Sayer et al., 2024; Delgadillo et al., 2018; Salyers et al., 2017).

Self-reported performance during assessments was also significantly higher when clinicians used the AI decision support tool, with the most notable improvements observed in their ability to prepare for assessments and complete them within the allotted appointment time. This may be because the AI tool provides substantial clinical information - much of which would otherwise need to be gathered during the assessment - as well as decision support that facilitates key clinical inferences such as diagnosis and treatment planning. By streamlining decision-making, the tool may free up time for interpersonal engagement between clinician and patient, particularly for rapport-building. Strengthening rapport, a core component of initial assessments, can be especially valuable in high-throughput settings such as NHS Talking Therapies, where limited appointment durations often constrain meaningful connection (De Geest & Meganck, 2019). Because the therapeutic relationship is a robust predictor of recovery and treatment adherence (Flückiger et al., 2018; Thompson & McCabe, 2012; Horvath et al., 2011), this enhancement could yield important downstream benefits for patients. Additionally, clinicians reported significantly lower cognitive load when supported by the AI tool. By reducing negative affect, mental demand, and effort - validated dimensions of cognitive load (Steel et al., 2015) - the tool may also serve as a protective factor against burnout.

These findings complement prior studies (Rollwage et al., 2023), showing that the benefits of AI-enabled decision support extend beyond the patient experience (e.g., with accessing mental health services) to also positively affecting clinicians. Specifically, prior work has demonstrated reduced therapy drop-out and faster recovery (Rollwage et al., 2024a, Rollwage et al., 2024b), while the present study provides new evidence of improved clinician wellbeing, enhanced task performance, and reduced cognitive load. This dual impact on both patients and clinicians highlights AI-powered front door technologies as tools that optimise patient pathways while safeguarding the workforce, addressing the intertwined challenges of patient recovery and clinician burnout in mental health services.

Our findings are of particular interest when considering that individual-level strategies (e.g., mindfulness, stress management) provide only modest and short-lived relief from clinician burnout, whereas systemic organisational interventions - particularly those addressing workload and workflow - are essential for sustained wellbeing (West et al., 2016; Panagioti et al., 2016; Awa et al., 2010). AI decision support tools represent such a systemic approach. By streamlining initial assessments and reducing task demands, they directly address the underlying drivers of burnout. In doing so, they target the foundational drivers of burnout, move beyond the limitations of reactive wellbeing programmes, and contribute to building a more resilient and sustainable mental health workforce.

Several limitations should be acknowledged. First, wellbeing and task performance were assessed using bespoke scales rather than validated instruments. This choice was made to ensure brevity and alignment with the specific context of NHS Talking Therapies, though it limits comparability with prior research. Future studies may replicate these findings using established or formally validated tools. Second, the reliance on self-report may have introduced unreliable estimates. Future work could extend these findings with objective cognitive load and performance measures made throughout the clinician working day (e.g., experience sampling methods, objective time tracking, or transcript analysis to measure clinician performance during assessments). Third, the self-reports were retrospective, requiring clinicians to recall and compare their experiences across different referral types, which may have introduced recall bias. Prospective or experimental designs would allow stronger causal inferences.

A further limitation of the present study is the observational nature of the patient assignment to different referral pathways (i.e. via the AI decision support tool and those referred through traditional pathways). Due to ethical considerations of freedom of patient choice, it was not possible to experimentally manipulate the availability of referral methods available to patients. Therefore, there may have been systematic differences in the patient populations referring through that AI tool or traditional referral pathways. Previous research investigating the impact of the same AI tool (Limbic Access) on patient outcomes, however, suggests no significant differences between these patient populations that could explain the observed effects (Rollwage et al., 2023). Nevertheless, future research could employ a randomised controlled design or combine clinician- and patient-level data to directly address this possibility.

## Conclusion

Our findings suggest that AI-powered decision support is an effective potential method for alleviating acute workforce pressures experienced by mental health practitioners in the UK, where nearly one-third of healthcare staff report time off for mental health reasons (Barnes, 2024), a striking indicator of systemic strain. Mental health services experience consistently higher turnover than sector-wide averages, as well as higher vacancy rates (Long et al., 2023), indicating that recruitment alone is not a sustainable solution. High task demands are strongly associated with impaired clinician wellbeing (Kim et al., 2018; Westwood et al., 2017; Steel et al., 2015) and reduced patient recovery rates (Sayer et al., 2024; Delgadillo et al., 2018; Salyers et al., 2017). Similar challenges are observed internationally, with burnout rates for US clinicians approaching 50% (Advisory Board, 2024), turnover in mental health roles as high as 22–35% annually (KLAS Research, 2024), and absences from work due mental health issues increasing by 300% from 2017 to 2023 (SHRM, 2024). Our findings therefore suggest that some of these pressures could be, at least in part, alleviated by supporting clinicians with intelligent digital support tools.

Such workforce pressures directly undermine service quality and continuity, creating a vicious cycle of burnout and inefficiency (Gilburt & Mallorie, 2024). Against this backdrop, scalable systemic interventions are urgently needed. Intelligent AI-powered tools, like the one studied here, offer a potential solution to cognitive and emotional strain and workflow inefficiencies. In doing so, such tools may help mitigate key drivers of staff turnover and contribute to a more resilient, sustainable mental health workforce, and eventually also benefit patient journeys to recovery.

## Data availability statement

Data required to reproduce the results of this study are available on GitHub at the following repository: https://github.com/LimbicAI/study-2025-clinician-preparedness.

## Acknowledgements

This work was supported by Limbic Limited.

## References

Advisory Board. (2024, January 31). Physician burnout and depression, in 5 charts. The Daily Briefing. https://www.advisory.com/daily-briefing/2024/01/31

Ashton-James, C. E., McNeilage, A. G., Avery, N. S., Robson, L. H. E., & Costa, D. (2021). Prevalence and predictors of burnout symptoms in multidisciplinary pain clinics: a mixed-methods study. Pain, 162(2), 503–513. 10.1097/j.pain.0000000000002042

Awa, W. L., Plaumann, M., & Walter, U. (2010). Burnout prevention: A review of intervention programs. Patient Education and Counseling, 78(2), 184–190. 10.1016/j.pec.2009.04.008

Barnes, A. (2024, April 7). Mental health absences adding to NHS staffing crisis | News, Press release | News. UNISON National. https://www.unison.org.uk/news/2024/04/mental-health-absences-adding-to-nhs-staffing-crisis/

De Geest, R. M., & Meganck, R. (2019). How Do Time Limits Affect Our Psychotherapies? A Literature Review. Psychologica Belgica, 59(1), 206–226. 10.5334/pb.475

Delgadillo, J., Saxon, D., & Barkham, M. (2018). Associations between therapists’ occupational burnout and their patients’ depression and anxiety treatment outcomes. Depression and Anxiety, 35(9), 844–850. 10.1002/da.22766

Devos, H., Gustafson, K., Ahmadnezhad, P., Liao, K., Mahnken, J. D., Brooks, W. M., & Burns, J. M. (2020). Psychometric Properties of NASA-TLX and Index of Cognitive Activity as Measures of Cognitive Workload in Older Adults. Brain Sciences, 10(12), 994. 10.3390/brainsci10120994

Flückiger, C., Del Re, A. C., Wampold, B. E., & Horvath, A. O. (2018). The alliance in adult psychotherapy: A meta-analytic synthesis. Psychotherapy, 55(4), 316–340. 10.1037/pst0000172

Gellert, G. A., Rasławska-Socha, J., Marcjasz, N., Price, T., Heyduk, A., Mlodawska, A., Kuszczyński, K., Jędruch, A., & Orzechowski, P. (2023). The Role of Virtual Triage in Improving Clinician Experience and Satisfaction: a Narrative Review. Telemedicine Reports, 4(1), 180–191. 10.1089/tmr.2023.0020

Gilburt, H., & Mallorie, S. (2024, February 21). Mental Health 360 | Workforce. The King’s Fund. https://www.kingsfund.org.uk/insight-and-analysis/long-reads/mental-health-360-workforce

Habicht, J., Viswanathan, S., Carrington, B., Hauser, T. U., Harper, R., & Rollwage, M. (2024). Closing the accessibility gap to mental health treatment with a personalized self-referral chatbot. Nature Medicine, 30. 10.1038/s41591-023-02766-x

Hart, S. G., & Staveland, L. E. (1988). Development of NASA-TLX (Task Load Index): Results of empirical and theoretical research. In Advances in psychology (Vol. 52, pp. 139–183). North-Holland.

Horvath, A. O., Del Re, A. C., Flückiger, C., & Symonds, D. (2011). Alliance in individual psychotherapy. Psychotherapy, 48(1), 9–16. 10.1037/a0022186

Kim, J. J., Brookman-Frazee, L., Gellatly, R., Stadnick, N., Barnett, M. L., & Lau, A. S. (2018). Predictors of burnout among community therapists in the sustainment phase of a system-driven implementation of multiple evidence-based practices in children’s mental health. Professional Psychology: Research and Practice, 49(2), 132–141. 10.1037/pro0000182

KLAS Research. (2024, December 3). Clinician Turnover 2024 - Arch Report. https://klasresearch.com/archcollaborative/report/clinician-turnover-2024/621

Kroenke, K., Spitzer, R. L., & Williams, J. B. W. (2001). The PHQ-9: Validity of a brief depression severity measure. Journal of General Internal Medicine, 16(9), 606–613.

Long, J., Ohlsen, S., Senek, M., Booth, A., Weich, S., & Wood, E. (2023). Realist synthesis of factors affecting retention of staff in UK adult mental health services. BMJ Open, 13(5), e070953–e070953. 10.1136/bmjopen-2022-070953

Meredith, L. S., Bouskill, K., Chang, J., Larkin, J., Motala, A., & Hempel, S. (2022). Predictors of burnout among US healthcare providers: a systematic review. BMJ Open, 12(8), e054243. 10.1136/bmjopen-2021-054243

NHS England. (2024, October 10). England’s NHS mental health services treat record 3.8 million people last year. NHS England. Retrieved 22/08/2025, from https://www.england.nhs.uk/2024/10/englands-nhs-mental-health-services-treat-record-3-8-million-people-last-year/

Owen, J., Crouch-Read, L., Smith, M., & Fisher, P. (2021). Stress and burnout in Improving Access to Psychological Therapies (IAPT) trainees: a systematic review. The Cognitive Behaviour Therapist, 14, e20. 10.1017/S1754470X21000179

Panagioti, M., Panagopoulou, E., Bower, P., Lewith, G., Kontopantelis, E., Chew-Graham, C., Dawson, S., van Marwijk, H., Geraghty, K., & Esmail, A. (2017). Controlled Interventions to Reduce Burnout in Physicians: A Systematic Review and Meta-analysis. JAMA Internal Medicine, 177(2), 195–205. 10.1001/jamainternmed.2016.7674

Rollwage, M., Habicht, J., Juechems, K., Carrington, B., Viswanathan, S., Stylianou, M., … & Harper, R. (2023). Using conversational AI to facilitate mental health assessments and improve clinical efficiency within psychotherapy services: real-world observational study. JMIR AI, 2(1), e44358.

Rollwage, M., Habicht, J., Juchems, K., Carrington, B., Hauser, T. U., & Harper, R. (2024a). Conversational AI facilitates mental health assessments and is associated with improved recovery rates. BMJ Innovations, 10(1–2), bmjinnov. 10.1136/bmjinnov-2023-001110

Rollwage, M., Juchems, K., Pisupati, S., Prichard, G., Balogh, A., McFadyen, J., … Harper, R. (2024b, November 27). The Limbic Layer: Transforming Large Language Models (LLMs) into Clinical Mental Health Experts. 10.31234/osf.io/9d7tp

Roy, A., Druker, S., Hoge, E. A., & Brewer, J. A. (2020). Physician Anxiety and Burnout: Symptom Correlates and a Prospective Pilot Study of App-Delivered Mindfulness Training. JMIR mHealth and uHealth, 8(4), e15608. 10.2196/15608

Salyers, M. P., Bonfils, K. A., Luther, L., Firmin, R. L., White, D. A., Adams, E. L., & Rollins, A. L. (2017). The Relationship Between Professional Burnout and Quality and Safety in Healthcare: A Meta-Analysis. Journal of General Internal Medicine, 32(4), 475–482. 10.1007/s11606-016-3886-9

Sayer, N. A., Kaplan, A., Nelson, D. B., Wiltsey Stirman, S., & Rosen, C. S. (2024). Clinician Burnout and Effectiveness of Guideline-Recommended Psychotherapies. JAMA Network Open, 7(4), e246858. 10.1001/jamanetworkopen.2024.6858

SHRM. (2024, March 24). Employee Leave Use Continues to Rise. TASB. https://www.tasb.org/news-insights/employee-leave-use-continues-to-rise

Spitzer, R. L., Kroenke, K., Williams, J. B. W., & Löwe, B. (2006). A brief measure for assessing generalized anxiety disorder: The GAD-7. Archives of Internal Medicine, 166(10), 1092–1097.

Steel, C., Macdonald, J., Schröder, T., & Mellor-Clark, J. (2015). Exhausted but not cynical: burnout in therapists working within Improving Access to Psychological Therapy Services. Journal of Mental Health, 24(1), 33–37. 10.3109/09638237.2014.971145

Thompson, L., & McCabe, R. (2012). The effect of clinician–patient alliance and communication on treatment adherence in mental health care: A systematic review. BMC Psychiatry, 12, 87. 10.1186/1471-244X-12-87

Turner, J., Knowles, E., Simpson, R., Sampson, F., Dixon, S., Long, J., … & Stone, T. (2021). Impact of NHS 111 Online on the NHS 111 telephone service and urgent care system: a mixed-methods study. Health Services and Delivery Research, 9(21).

West, C. P., Dyrbye, L. N., Erwin, P. J., & Shanafelt, T. D. (2016). Interventions to prevent and reduce physician burnout: a systematic review and meta-analysis. The Lancet, 388(10057), 2272–2281. 10.1016/s0140-6736(16)31279-x

West Suffolk NHS Foundation Trust. (n.d.). After the Academy: A guide to future careers [PDF]. https://www.wsh.nhs.uk/Join-our-team/Health-and-care-academies/docs/After-the-academy.pdf

Westwood, S., Morison, L., Allt, J., & Holmes, N. (2017). Predictors of emotional exhaustion, disengagement and burnout among improving access to psychological therapies (IAPT) practitioners. Journal of Mental Health (Abingdon, England), 26(2), 172–179. 10.1080/09638237.2016.1276540

Willman, A. S. (2023). Evaluation of eConsult use by Defence Primary Healthcare primary care clinicians using a mixed-method approach. BMJ Military Health, 169(e1), e39–e43. 10.1136/bmjmilitary-2020-001660

Xiao, Y. M., Wang, Z. M., Wang, M. Z., & Lan, Y. J. (2005). The appraisal of reliability and validity of subjective workload assessment technique and NASA-task load index. Chinese journal of industrial hygiene and occupational diseases, 23(3), 178–181.

