## Supplementary Materials for "AI-driven Mental Health Decision Support Enhances Clinician Resilience and Preparedness"

### Outcome measures

#### Wellbeing

Questions (scale from 1 [not at all] to 7 [extremely]):

- **Energy**: "How energised do you feel about these assessments?"
- **Confidence**: "How confident do you feel about performing these assessments?"
- **Comfort**: "How comfortable do you feel about conducting these assessments?"
- **Stress**: "How stressed do you feel about these assessments?"
- **Uncertainty**: "How uncertain do you feel about doing these assessments?"
- **Anxiety**: "How anxious do you feel before these assessments?"

Negative items (anxiety, uncertainty, stress) have been reverse-scored.

#### Task Performance

Questions (scale from 1 [very difficult] to 7 [very easy]):

- **Prepare Assessment**: "How easy is it to prepare for the assessment?"
- **Determine Treatment**: "How easy is it to determine the appropriate treatment?"
- **Risk Assessment**: "How easy is it to conduct a risk assessment?"
- **Identify Diagnosis**: "How easy is it to identify a diagnosis?"
- **Time Limit**: "How easy is it to complete the assessment within your service’s time limit?"
- **Build Relationship**: "How easy is it to build a relationship with the patient?"
- **Manage Expectations**: "How easy is it to manage a patient’s expectations for their diagnosis and/or treatment plan?"

#### NASA Task Index

Questions (scale from 1 [not at all] to 7 [extremely]):

- **Mental Demand**: "How mentally demanding is the assessment?"
- **Hurried Pace**: "How hurried or rushed is the pace of the assessment?"
- **Success / Accomplishment**: "How successful are you in accomplishing what you want to accomplish?"
- **Work Hard**: "How hard do you have to work to accomplish your level of performance?"
- **Negative Feelings**: "How insecure, discouraged, irritated, stressed, or annoyed do you feel?"

The “Success / Accomplishment” item is reverse-scored.
